# The validation of the original and modified Caprini score in COVID-19 patients

**DOI:** 10.1101/2020.06.22.20137075

**Authors:** Sergey Tsaplin, Ilya Schastlivtsev, Kirill Lobastov, Sergey Zhuravlev, Victor Barinov, Joseph A Caprini

**Author notes:** **Corresponding author and the post-publication corresponding author**: Kirill Lobastov, 117997, Ostrovitianov street build. 1, Department of General Surgery and Radiology, Moscow, Russian Federation.

## Abstract

**Objective:** The study aimed to validate the original Caprini score and its modifications considering coronavirus disease (COVID-19) as a severe prothrombotic condition in patients admitted to the hospital with confirmed infection.

**Methods:** The relevant data were extracted from the electronic medical records with the implemented Caprini score and were evaluated retrospectively. The score was calculated twice: by the physician at the admission and by the investigator at discharge or after death. The second calculation at discharge, considered additional risk factors that occurred during inpatient treatment. Besides the original Caprini score (a version of 2005), the modified version added the elevation of D-dimer and specific scores for COVID-19 as follows: 2 points for asymptomatic, 3 points for symptomatic and 5 points for symptomatic infection with positive D-dimer, were evaluated in a retrospective manner. The primary endpoint was symptomatic venous thromboembolism (VTE) confirmed by appropriate imaging testing or dissection. The secondary endpoint included the unfavorable outcome as a combination of symptomatic VTE, admission to the intensive care unit, the requirement for invasive mechanical ventilation, and death. The association of eight different versions of the Caprini score with outcomes was evaluated.

**Results:** Totally 168 patients (83 males and 85 females at the age of 58.3±12.7 years old) were admitted to the hospital between April 30 and May 29, 2020, and were discharged or died up to the time of data analysis. The original Caprini score varied between 2-12 (5.4±1.8) at the admission and between 2-15 (5.9±2.5) at discharge or death. The presence of the virus increased these scores and resulted in an increased score with the maximal value for those including COVID-19 points (10.0±3.0). Patients received prophylactic (2.4%), intermediate (76.8%), or therapeutic (20.8%) doses of enoxaparin. Despite this, the symptomatic VTE was detected in 11 (6.5%) and unfavorable outcomes in 31 (18.5%) patients. The Caprini score of all eight versions demonstrated a significant association with VTE with the highest predictability for the original scale when assessed at discharge. Supplementation of the original score by elevated D-dimer improved predictability only at the admission. Four versions of the Caprini score calculated at the admission had a significant correlation with the unfavorable outcome with the minor advantages of specific COVID-19 points.

**Conclusion:** The study identified a significant correlation between the Caprini score and the risk of VTE or unfavorable outcomes in COVID-19 patients. All models, including specific COVID-19 scores, showed high predictability with minor differences.

**ARTICLE HIGHLIGHTS:** *Type of Research:* A single-center retrospective analysis of prospectively collected data.

*Key Findings:* The original version of the Caprini score and its modifications considering the elevation of D-dimer and specific COVID-19 points demonstrated a significant association with symptomatic VTE and unfavorable outcome in 168 hospitalized COVID-19 patients, of whom 6.5% developed symptomatic VTE and 18.5% - unfavorable outcome despite routine pharmacoprophylaxis.

*Take Home Message:* The Caprini score allows stratification of COVID-19 inpatients according to their VTE risk and identification of subjects at extremely high risk.

**TABLE OF CONTENTS SUMMARY:** This retrospective analysis of prospectively collected data demonstrates the significant association between the original and modified Caprini score and symptomatic VTE or unfavorable outcome in 168 patients with confirmed COVID-19. The Caprini score may be used for VTE risk assessment, and identification of persons at extremely high risk among COVID-19 patients admitted to the hospital.

## Introduction

Coronavirus disease (COVID-19) is a highly infectious disease caused by the SARS-CoV-2 virus leading to the development of severe pneumonia and acute respiratory distress syndrome (ARDS). The infection has appeared in Wuhan, Hubei Province, China, in December of 2019, and has been spread globally.^1^ The high prevalence of venous thromboembolism (VTE), including deep vein thrombosis (DVT), pulmonary embolism (PE), and pulmonary artery (PA) thrombosis, among the inpatients and critically ill patients, have been reported since the beginning of the pandemic.^2-11^ The overall duplex ultrasound scan (DUS) detected the presence of DVT in 46% of the patients in the general ward and up to 79% of the patients in the intensive care unit (ICU).^10, 11^ Proven PE using contrast computed tomography (CT) scan was observed in 30% of all COVID-19 patients.^8^ The evidence of small and mid-sized PA thrombosis and microthrombi in alveolar capillaries was found in most of the deceased patients in parallel with the occlusion of big branches of PA in 9-33% of dissections.^12-16^ Considering the high incidence of thrombotic complications in COVID-19, most of the current guidelines suggest routine pahrmacoprophylaxis using low-molecular-weight heparin (LMWH) or unfractionated heparin (UFH) for patients admitted to the hospital.^17-21^ Some publications support the intensification of anticoagulation (intermediate to therapeutic doses of heparin) in subjects at individually highest risk for VTE including critical illness, obesity, and high level of D-dimer.^17, 19, 20^ The tools mentioned in these publications for VTE risk assessment include: the Caprini score, Padua score, and IMPROVE VTE score.^18-20^ The Padua score was assessed in two papers without a clear correlation with VTE events.^22, 23^ To the best of our knowledge, the Caprini and IMPROVE VTE scores have not been validated in COVID-19 patients.

To cover this gap, we decided to perform the current study aimed to validate the original Caprini score and its modified version considering COVID-19 as a severe prothrombotic condition in patients admitted to the hospital with confirmed SARS-CoV-2 infection. We also tested the influence of several levels of D-dimer combined with the original Caprini score to improve the identification of patients at the highest risk of VTE.

## Methods

This study is a single-center retrospective analysis of prospectively collected data obtained from medical records of patients admitted at the Clinical Hospital no.1 (Volynskaya) of the President’s Administration of the Russian Federation (Moscow, Russian Federation) with verified COVID-19. The hospital stopped standard medical care and was reoriented for COVID-19 on April 30, 2020. Two years before the Caprini score (a version of 2005 in Russian translation^24^) was integrated into the electronic medical records (EMR) and became mandatory for assessment. The EMRs were extracted on May 30, 2020, when all COVID-19 patients were discharged, and the hospital turned back to the standard care.

The initial Caprini score was calculated by the physician at the time of patients admission. It did not take into account some risk factors related to inpatient treatment (bed rest for >72 hours, central venous catheter, acquired thrombophilia, etc.). Standard coagulation testing, including the level of D-dimer, prothrombin time (PT), activated partial thromboplastin time (APTT), was performed in all admitted patients but was not considered within the primary assessment of Caprini score. The reason being at the time of admission, the blood test results were not known by the initial examiner. In Russia, the indication for inpatient treatment for adults is a moderate to severe disease according to the locally adopted criteria with respiratory rate >22 per min or SpO_2_ <93% with room air, a mild illness in subjects over 65 years old with significant co-morbidities (chronic heart failure, diabetes mellitus, chronic obstructive pulmonary disease [COPD]), and pregnancy. We decided that all patients with moderate to severe illness and/or representing specific signs of pneumonia on a chest CT scan should receive one Caprini point for pneumonia at the admission as well as one additional score for medical illness with bed rest (Table I).

**Table I.** The distribution of the individual risk factors for VTE according to the Caprini score.

| **Risk factor** | **Score** | **Number of patients (%)** |
| --- | --- | --- |
| Age 41-60 years | 1 | 75 (45%) |
| Swollen legs | 1 | 32 (19%) |
| Varicose veins | 1 | 32 (19%) |
| Obesity (BMI >25) | 1 | 131 (78%) |
| Minor surgery | 1 | 0 (0%) |
| Sepsis (<1 month) | 1 | 72 (43%) |
| Acute miocardial infarction | 1 | 0 (0%) |
| Congestive heart failure (<1 month) | 1 | 24 (14%) |
| Medical patient currently at bed rest | 1 | 168 (100%) |
| History of inflammatory bowel disease | 1 | 4 (2%) |
| History of prior major surgery (<1 month) | 1 | 0 (0.0%) |
| Abnormal pulmonary function (COPD) | 1 | 8 (5%) |
| Serious lung disease including pneumonia (<1 month) | 1 | 168 (100%) |
| Oral contraceptives or hormone replacement therapy | 1 | 0 (0%) |
| Pregnancy or postpartum (< 1 month) | 1 | 0 (0%) |
| History of unexplained stillborn infant, recurrent spontaneous abortion ($\geq 3$), premature birth with toxemia or growth-restricted infant | 1 | 1 (0.6%) |
| Age 61-74 years | 2 | 61 (36%) |
| Central venous access<sup>†</sup> | 2 | 22 (13%) |
| Arthroscopic surgery | 2 | 0 (0%) |
| Major surgery (>45 min) | 2 | 0 (0%) |
| Malignancy (present or previous) | 2 | 0 (0%) |
| Laparoscopic surgery (>45 min) | 2 | 0 (0%) |
| Patient confined to bed (>72 hours)<sup>‡</sup> | 2 | 14 (8%) |
| Immobilizing plaster cast (<1 month) | 2 | 0 (0%) |
| COVID-19 asymptomatic<sup>□</sup> | 2 | 0 (0%) |
| Age 75 years or older | 3 | 19 (11%) |
| History of DVT/PE | 3 | 6 (4%) |
| Family history of thrombosis | 3 | 0 (0%) |
| Positive factor V Leiden | 3 | 0 (0%) |
| Positive prothrombin 20210A | 3 | 0 (0%) |
| Elevated serum homocysteine | 3 | 0 (0%) |
| Heparin-induced thrombocytopenia | 3 | 0 (0%) |
| Positive lupus anticoagulant | 3 | 0 (0%) |
| Elevated anti-cardiolipin antibodies | 3 | 0 (0%) |
| Other congenital or acquired thrombophilia: D-dimer >ULN<sup>□</sup> | 3 | 102 (61%) |
| Other congenital or acquired thrombophilia: D-dimer >3ULN<sup>□</sup> | 3 | 15 (9%) |
| COVID-19 symptomatic<sup>□</sup> | 3 | 66 (39%) |
| Stroke (<1 month) | 5 | 0 (0%) |
| Multiple trauma (<1 month) | 5 | 0 (0%) |
| Elective major lower extremity arthroplasty | 5 | 0 (0%) |
| Hip, pelvis or leg fracture (<1 month) | 5 | 0 (0%) |
| Acute spinal cord injury, paralysis (<1 month) | 5 | 0 (0%) |
| COVID-19 symptomatic with positive D-dimer<sup>□</sup> | 5 | 102 (61%) |

<sup>†</sup>Evaluated at discharge or after death
<sup>□</sup>Evaluated within the modified versions of Caprini score
BMI – body mass index; COPD – chronic obstructive pulmonary disease; DVT – deep vein thrombosis; PE – pulmonary embolism; ULN – upper limit or normal

The final score was assessed by the two investigators via EMR after patients’ discharge or death. The additional risk factors related to inpatient treatment were evaluated. Also, the elevated D-dimer at admission was assessed as “acquired” thrombophilia from the block of 3 scores. We used a liberal or strict approach in the assessment of D-dimer and achieved two different versions of the Caprini score. In the liberal version, we put an additional 3 points for any patient who had elevated D-dimer over the upper limit of normal (ULN: >0,55 mg/L) at the admission. Within the strict version, only patients with D-dimer >3 times ULN (>1,5 mg/L) received the additional 3 points. Due to the coronavirus pandemic, one of the authors (JA Caprini) proposed using a modified score for COVID-19 patients. It adds 2 points for COVID-19 patients with asymptomatic disease, 3 points for those with symptomatic illness, and 5 points for symptomatic patients with a positive D-dimer. This modification also was used for the retrospective analysis.

We calculated the four different Caprini scores obtained at the admission and the discharge or death. We achieved eight versions of the scale: Caprini[orig:adm] - original score calculated at admission; Caprini[orig:fin] - original score calculated at discharge or death; Caprini[Dd>ULN:adm] - score with liberal D-dimer calculated at admission; Caprini[Dd>ULN:fin] - score with liberal D-dimer calculated at discharge or death; Caprini[Dd>3ULN:adm] - score with strict D-dimer calculated at admission; Caprini[Dd>3ULN:fin] - score with strict D-dimer calculated at discharge or death; Caprini[COVID-19:adm] – modified score calculated at admission; Caprini[COVID-19:fin] – modified score calculated at discharge or death.

The standard prevention of VTE included prophylactic or intermediate doses of LMWH. The local protocol proposed in our clinic required intermediate doses of LMWH for all patients without bleeding risk. It was suggested at the background of the high prevalence of VTE and PA thrombosis in COVID-19 patients. Although not all international guidelines support this approach, it was not contradictory to the interim Russian national recommendations. Only patients at high risk of bleeding were managed with the standard prophylactic doses LMWH. The therapeutic anticoagulation was prescribed for subjects without confirmed VTE in case of a critical level of D-dimer (>6 times the ULN) or suspected thrombi in PA (e.g., hypoxemia disproportionate to known respiratory pathologies, or acute unexplained right ventricular dysfunction). The local protocol suggested empiric initiation of therapeutic anticoagulation in case of presumptive VTE with the mandatory further verification by appropriate imaging tests before discharge. A duplex ultrasound scan was used to assess superficial and deep veins of lower and upper extremities, computed tomography pulmonary angiogram (CTPA) - to exclude thrombi in the pulmonary artery. If the patient died, the autopsy with an evaluation of lung vessels and veins of lower limbs was obligatory for the final VTE verification.

The primary endpoint was symptomatic venous thromboembolism confirmed by appropriate imaging testing or dissection at autopsy. The secondary endpoints included: (1) the unfavorable outcome as a combination of symptomatic VTE, admission to the ICU, the requirement for invasive mechanical ventilation, and death, and (2) a bleeding outcome as a combination of major and clinically relevant non-major (CRNM) bleeding by the criteria of International Society of Thrombosis and Hemostasis.^25, 26^ All events were assessed in a retrospective manner by three experienced in vascular surgery investigators, according to the data extracted from EMRs. The correlation with the VTE event was evaluated for the Caprini score calculated either at admission or discharge or death. In contrast, the correlation between an unfavorable outcome and the Caprini score was done only at the time of admission.

No need for ethical approval and a waiver of informed consent because of the retrospective character of the analysis in the urgent situation was confirmed by the Institutional Review Board of Clinical Hospital no.1 of the President’s Administration of the Russian Federation.

Statistical analysis. All absolute values are represented by mean with standard deviation (SD) in the normal distribution or by a median with interquartile range (IQR) 25-75 percentile in the non-normal distribution assessed by Kolmogorov-Smirnov test. The relative values are represented as percents, with a 95% confidence interval (CI) calculated by Wilson. The data were compared by paired t-test or ANOVA. The association between the Caprini score and outcomes was evaluated by the generalized linear model (GLM), a form of linear regression analysis. The predictability of Caprini scores for proposed outcomes was assessed by a single-factor univariate logistic regression analysis and evaluation of receiver operating characteristic (ROC) curves with their areas under the curve (AUC). The cut-off for the Caprini score with current sensitivity and specificity was extracted from the coordinates of ROC curves. The analysis was performed using SPSS version 26 software package (IBM Corp, Armonk, NY). The value of p<0.05 was considered statistically significant.

## Results

A total of 168 patients were admitted to the hospital between April 30 and May 29, 2020. Up to the time of data extraction, everybody was discharged or died, and every EMR was eligible for analysis. The number of males and females was equivalent (83 and 85, respectively). The age varied between 21 and 86 (mean of 58.3±12.7) years old. COVID-19 diagnosis was confirmed with polymerase chain reaction on nasopharyngeal swabs of 140 patients (83%); 28 (17%) despite the negative result had a typical pattern of viral pneumonia on chest CT scan (ground-glass opacity) as assessed by the experienced radiologists. The interim Russian national guidelines classified the disease severity as moderate (body temperature >38 degrees of Celsium, respiratory rate >22 per min, dyspnea in physical activity, pneumonia by chest CT scan, SpO_2_<95%, C-reactive protein >10 mg/mL) in 155 (92%) patients. Severe disease criteria (respiratory rate >30 per min, SpO_2_≤93%, PaO_2_ /FiO_2_ ≤300 mm Hg, disease progression by chest CT scan, a decrease of consciousness or agitation, unstable hemodynamics, serum lactate >2 mmol/L, qSOFA >2) were found in 13 (8%) subjects. Time to admission from the onset of symptoms varied between 1 and 24 (mean of 7.2±4.0) days. During this period, patients were treated at home with symptomatic therapy, chloroquine or hydroxychloroquine, and antibiotics.

The PT at the admission ranged from 9.3 to 29.1 sec (mean of 11.6±1.9 sec; the normal range of 9.8-12.1 sec). It was prolonged in nine (5%) patients. The APTT varied between 22.0-54.1 sec (mean of 32.9±5.0 sec; the normal range of 26.4-37.5 sec). It was increased in 44 (26%) subjects. The D-dimer varied from 0,1 to 6.7 mg/L (median of 0.6 mg/L with the IQR of 0.4-1.0 mg/L; ULN is 0.55 mg/L). Any elevation of D-dimer was observed in 102 (61%) patients and elevation >3 times ULN – in 15 (9%) subjects. We did not use the age-adjusted ULN for D-dimer like all previous reports on its level in patients with COVID-19.^3, 8, 10, 27-34^

All patients received therapy with chloroquine or hydroxychloroquine, antibiotics, non-invasive and invasive oxygen therapy, symptomatic treatment, and prophylactic anticoagulation. The LMWH was used in the standard prophylactic dose (enoxaparin 40 mg once daily) in 4 (2.4%) patients, in the intermediate dose (enoxaparin 60-80 mg once daily) in 129 (76.8%) patients, and in therapeutic doses (enoxaparin 1 mg/kg twice daily) in 35 (20.8%) patients. The duration of inpatient treatment varied from 1 to 29 days (mean of 13.9±4.0 days).

The risk factor distribution by the Caprini score is represented in Table I. By the original scale, the score ranged from 2 to 12 (mean of 5.4±1.8) as calculated at the admission and between 2 and 15 (mean of 5.9±2.5) as calculated at discharge or death. Among all modifications, the highest score was observed with Caprini[COVID-19] version followed by Caprini[Dd>ULN], Caprini[Dd>3ULN], and the original version. All scores significantly increased when they were measured at the end of hospitalization (Table II).

**Table II.**
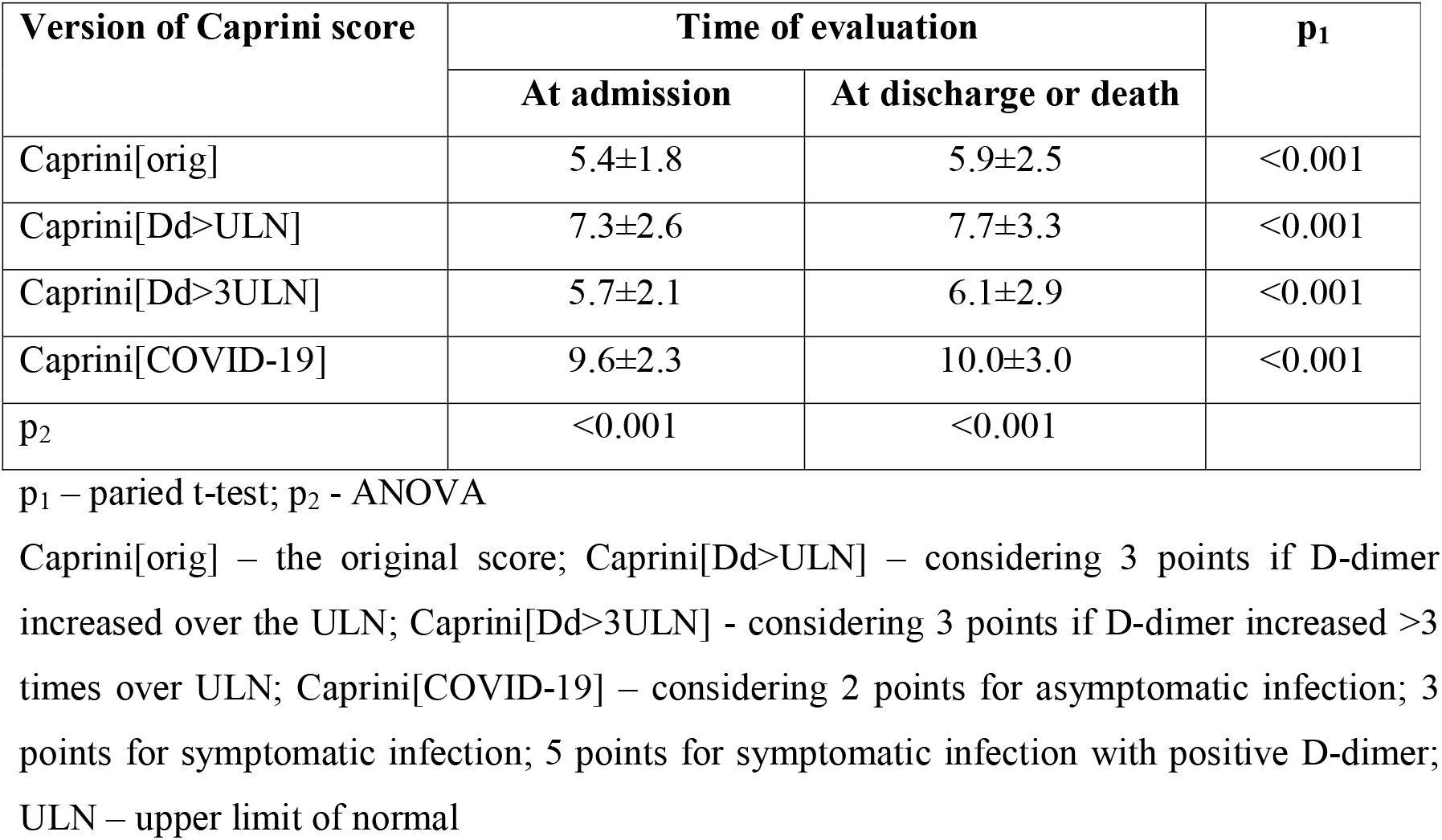
The value of Caprini score according to the version and time of evaluation.

The symptomatic VTE was detected in 11 of 168 (6.5%; 95% CI, 3.7-11.3%) patients including catheter-related venous thrombosis of upper limbs in 3 (1.8%; 95% CI, 0.6-5.1%) cases; isolated DVT in 2 (1.2%; 95% CI, 0.3-4.3%) cases; combination of DVT and PE in 1 (0.6%; 95% CI, 0.1-3.3%) case, and isolated PE in 5 (3.0%; 95% CI, 1.3-6.8%) cases. Of three DVT, two affected the distal veins, and one – the proximal veins. Thrombosis of calf muscle veins was the only one of these leg clots associated with PE. Of six pulmonary embolism, five were fatal and confirmed by autopsy; the last one was confirmed by CTPA. All lesions affected segmental and subsegmental branches of PA. All VTE events developed despite intermediate (n=5) or therapeutic (n=6) doses of LMWH.

The unfavorable outcome was detected in 31 (18.5%; 95% CI, 13.1-24.7%) patients including admission to the ICU in 28 (16.7%; 95% CI, 11.8-23.1%), the requirement for mechanical ventilation in 8 (4.8%; 95% CI, 2.5-9.2%), and death in 8 (4.8%; 95% CI, 2.5-9.2%) cases. Of 8 patients on mechanical ventilation, 7 (88%) finally died. The CRNM bleeding was detected in only 2 (1.2%; 95% CI, 0.3-4.3%) subjects represented by subcutaneous hematoma required surgical drainage and uterine bleeding. Both occurred while receiving a therapeutic and intermediate dose of LMWH, respectively.

The statistically significant association with symptomatic VTE was observed for all versions of the Caprini score calculated either at the admission or discharge or death (p<0.001, Figures 1 and 2). The high predictability was confirmed by the single-factor logistic regression analysis (Table III) and the analysis of ROC curves (Table IV, Figure 4). Among the scores calculated at the admission, the highest predictability by the area under the curve was found for the version considering any elevation of D-dimer (Caprini[Dd>ULN:adm]). The score of ≥10 with the sensitivity of 73% and specificity of 85% predicted the occurrence of symptomatic venous thromboembolism. However, the standard version (Caprini[orig:adm]) demonstrated high predictability as well: with a sensitivity of 73% and the specificity of 80%, the score of ≥7 predicted VTE. Among those scores obtained at the discharge or death, the best predictability was observed for the original version of the scale (Caprini[orig:fin]): with the sensitivity of 73% and specificity of 96% score of ≥11 predicted VTE. Also, this model had the highest AUC among all evaluated. The second highest predictability was observed for the version considering any elevation of D-dimer (Caprini[Dd>ULN:fin]): with the sensitivity of 73% and specificity of 97% score of ≥14 predicted VTE. Generally, better predictability and higher specificity were observed for the Caprini score obtained at the end of inpatient treatment.

**Table III.** Predictability of different versions of Caprini score for symptomatic VTE and unfavorable outcome in COVID-19 patients by the single-factor logistic regression.

| Version of Caprini score | OR | 95% CI | p |
| --- | --- | --- | --- |
| **Symptomatic VTE** |  |  |  |
| Caprini[orig:adm] | 1.87 | 1.34-2.61 | <0.001 |
| Caprini[Dd>ULN:adm] | 1.63 | 1.23-2.17 | 0.001 |
| Caprini[Dd>3ULN:adm] | 1.59 | 1.24-2.05 | <0.001 |
| Caprini[COVID-19:adm] | 1.73 | 1.27-2.34 | <0.001 |
| Caprini[orig:fin] | 1.73 | 1.36-2.20 | <0.001 |
| Caprini[Dd>ULN:fin] | 1.58 | 1.28-1.95 | <0.001 |
| Caprini[Dd>3ULN:fin] | 1.61 | 1.30-2.00 | <0.001 |
| Caprini[COVID-19:fin] | 1.63 | 1.31-2.03 | <0.001 |
| **Unfavorable outcome** |  |  |  |
| Caprini[orig:adm] | 1.84 | 1.43-2.38 | <0.001 |
| Caprini[Dd>ULN:adm] | 1.57 | 1.29-1.90 | <0.001 |
| Caprini[Dd>3ULN:adm] | 1.61 | 1.31-1.98 | <0.001 |
| Caprini[COVID-19:adm] | 1.68 | 1.35-2.10 | <0.001 |

Unfavorable outcome represents the combination of symptomatic VTE, admission to the ICU, requirement for mechanical ventilation, and death.

**Table IV.** Predictability of different versions of Caprini score for symptomatic VTE and unfavorable outcome in COVID-19 patients by the analysis of ROC curve.

| Version of Caprini score | AUC±SD | p | Cut-off | Sensitivity | Specificity |
| --- | --- | --- | --- | --- | --- |
| **Symptomatic VTE** |  |  |  |  |  |
| Caprini[orig:adm] | 0.769±0.090 | 0.003 | 7 | 73% | 80% |
| Caprini[Dd>ULN:adm] | 0.776±0.096 | 0.002 | 10 | 73% | 85% |
| Caprini[Dd>3ULN:adm] | 0.757±0.090 | 0.004 | 7 | 73% | 76% |
| Caprini[COVID-19:adm] | 0.772±0.097 | 0.003 | 12 | 73% | 84% |
| Caprini[orig:fin] | 0.803±0.095 | 0.001 | 11 | 73% | 96% |
| Caprini[Dd>ULN:fin] | 0.801±0.100 | 0.001 | 14 | 73% | 97% |
| Caprini[Dd>3ULN:fin] | 0.797±0.096 | 0.001 | 11 | 73% | 94% |
| Caprini[COVID-19:fin] | 0.799±0.100 | 0.001 | 16 | 73% | 97% |
| **Unfavorable outcome** |  |  |  |  |  |
| Caprini[orig:adm] | 0.775±0.046 | <0.001 | 6 | 74% | 66% |
| Caprini[Dd>ULN:adm] | 0.775±0.049 | <0.001 | 9 | 68% | 75% |
| Caprini[Dd>3ULN:adm] | 0.778±0.046 | <0.001 | 6 | 77% | 63% |
| Caprini[COVID-19:adm] | 0.780±0.048 | <0.001 | 11 | 68% | 75% |

Unfavorable outcome represents the combination of symptomatic VTE, admission to the ICU, requirement for mechanical ventilation, and death.

**Figure 1.**
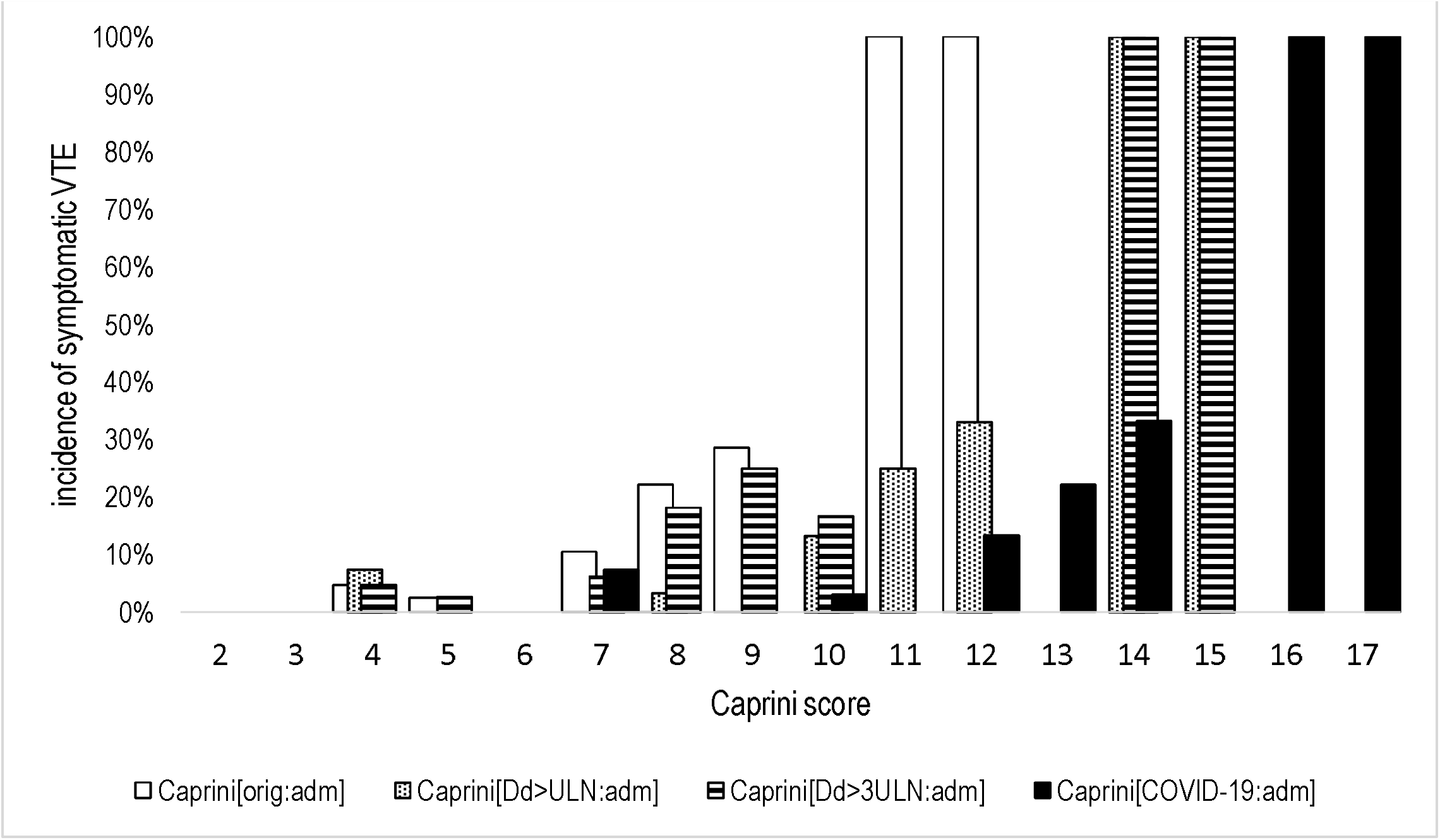
The incidence of VTE according to the different modifications of Caprini score assessed at the time of admission. Caprini[orig:adm] – the original score assessed at admission (p<0.001 by GLM); Caprini[Dd>ULN:adm] - considering 3 points if D-dimer increased over ULN, score assessed at admission (p<0.001 by GLM); Caprini[Dd>3ULN:adm] - considering 3 points if D-dimer increased >3 times over ULN, score assessed at admission (p<0.001 by GLM); Caprini[COVID-19:adm] – considering 2 points for asymptomatic infection, 3 points for symptomatic infection, 5 points for symptomatic infection with positive D-dimer, score assessed at admission (p<0.001 by GLM); VTE – venous thromboembolism; ULN – upper limit of normal

**Figure 2.**
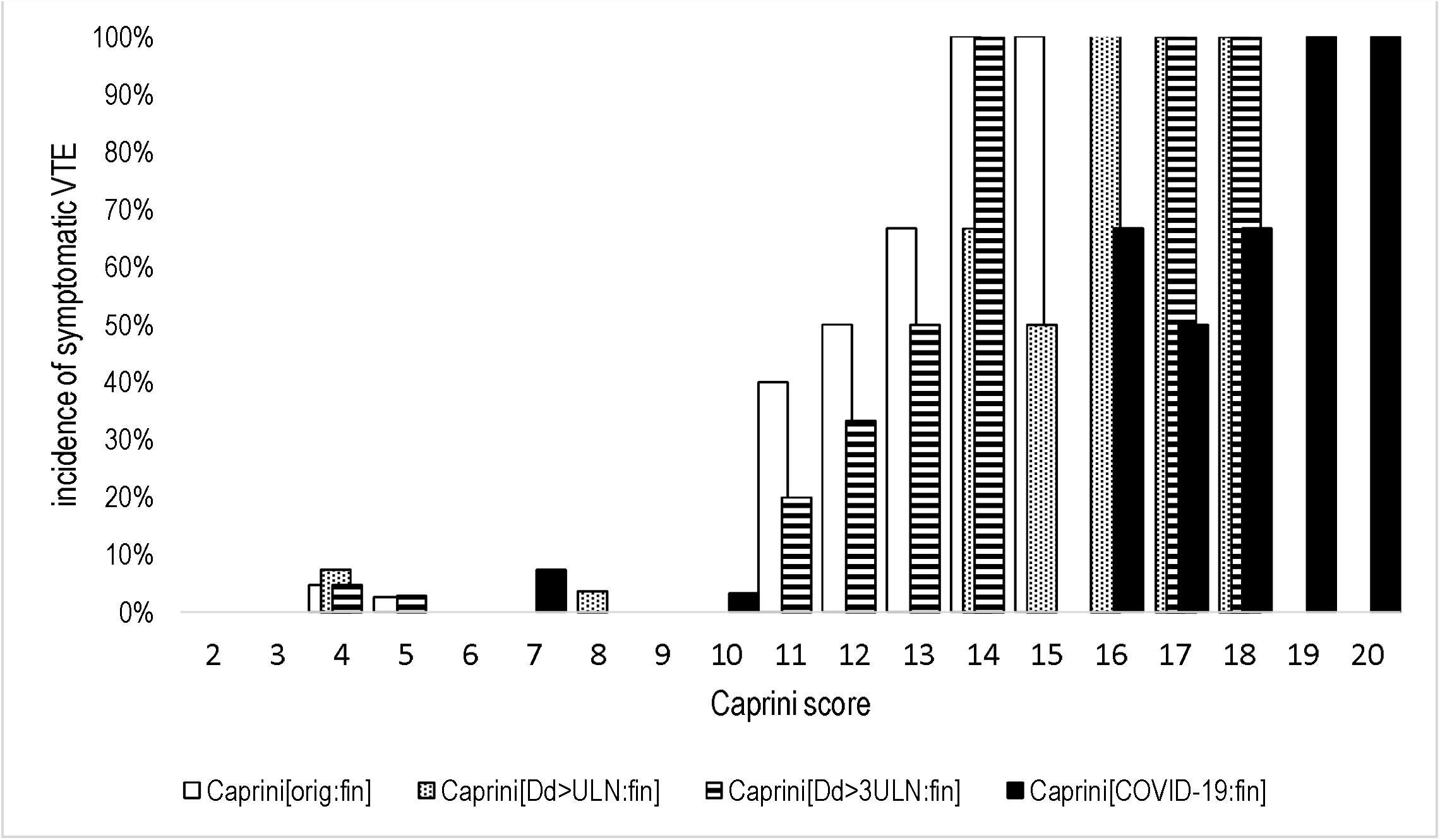
The incidence of symptomatic VTE according to the different modifications of Caprini score assessed at the time of discharge or death. Caprini[orig:fin] – the original score assessed at discharge or death (p<0.001 by GLM); Caprini[Dd>ULN:fin] - considering 3 points if D-dimer increased over ULN, score assessed at discharge or death (p<0.001 by GLM); Caprini[Dd>3ULN:fin] - considering 3 points if D-dimer increased >3 times over ULN, score assessed at discharge or death (p<0.001 by GLM); Caprini[COVID-19:fin] – considering 2 points for asymptomatic infection, 3 points for symptomatic infection, 5 points for symptomatic infection with positive D-dimer, score assessed at discharge or death (p<0.001 by GLM); VTE – venous thromboembolism; ULN – upper limit of normal

**Figure 3.**
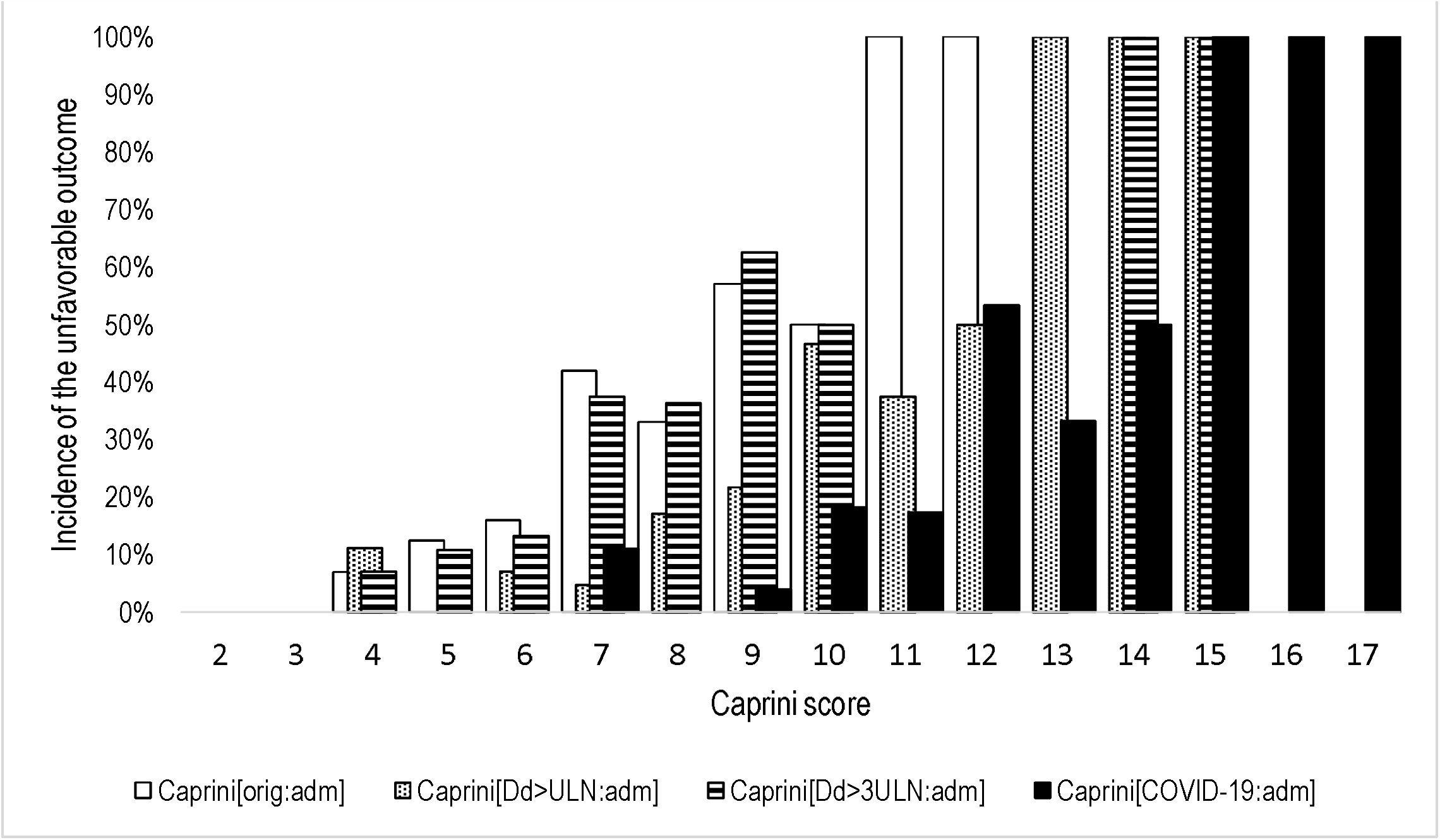
The incidence of unfavorable outcome according to the different modifications of Caprini score assessed at the time of admission. Unfavorable outcome combined symptomatic VTE, admission to the intensive care unit, the requirement for mechanical ventilation and death Caprini[orig:adm] – the original score assessed at admission (p<0.001 by GLM); Caprini[Dd>ULN:adm] - considering 3 points if D-dimer increased over ULN, score assessed at admission (p<0.001 by GLM); Caprini[Dd>3ULN:adm] - considering 3 points if D-dimer increased >3 times over ULN, score assessed at admission (p<0.001 by GLM); Caprini[COVID-19:adm] – considering 2 points for asymptomatic infection, 3 points for symptomatic infection, 5 points for symptomatic infection with positive D-dimer, score assessed at admission (p<0.001 by GLM); ULN – upper limit of normal

**Figure 4.**
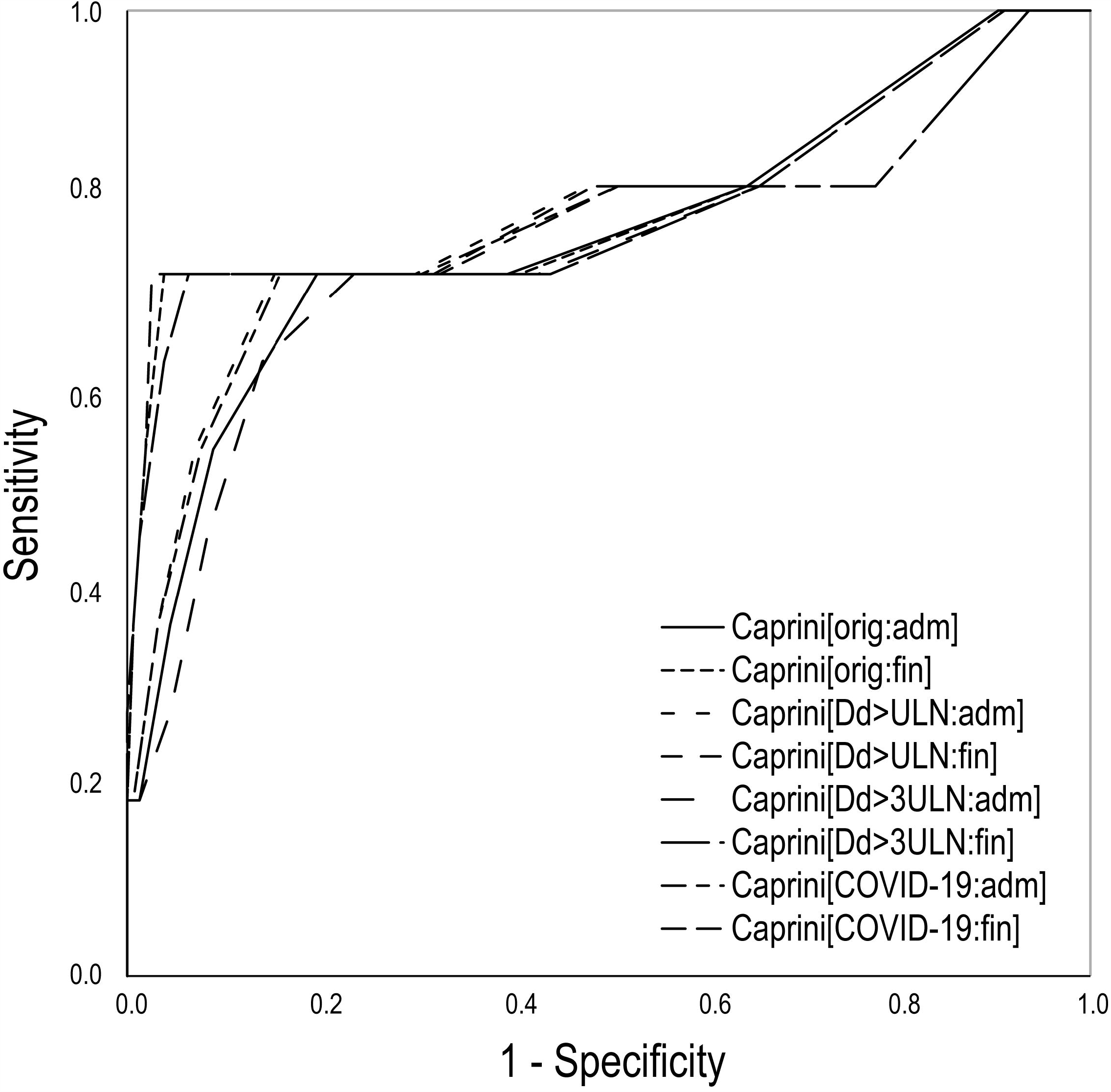
Predictability of different versions of Caprini score for symptomatic VTE by the ROC curves. Caprini[orig:adm] – the original score assessed at admission; Caprini[orig:fin] – the original score assessed at discharge or death; Caprini[Dd>ULN:adm] – considering 3 points if D-dimer increased over ULN, score assessed at admission; Caprini[Dd>ULN:fin] – considering 3 points if D-dimer increased over ULN, score assessed at discharge or death; Caprini[Dd>3ULN:adm] - considering 3 points if D-dimer increased >3 times over ULN, score assessed at admission; Caprini[Dd>3ULN:fin] - considering 3 points if D-dimer increased >3 times over ULN, score assessed at discharge or death; Caprini[COVID-19:adm] - considering 2 points for asymptomatic infection, 3 points for symptomatic infection, 5 points for symptomatic infection with positive D-dimer, score assessed at admission; Caprini[COVID-19:fin] – considering 2 points for asymptomatic infection, 3 points for symptomatic infection, 5 points for symptomatic infection with positive D-dimer, score assessed at discharge or death; ROC – receiver operating characteristic

The significant association between the Caprini score of all versions obtained at the admission and the unfavorable outcome in COVID-19 patients was observed (Figure 3). The logistic regression and the analysis of ROC curves confirmed the high predictability of all models (Table III, IV, Figure 5). The modified version considering the COVID-19 points (Caprini[COVID-19:adm]) provided the highest AUC. The score of ≥11 with the sensitivity of 68% and specificity of 75% predicted the unfavorable outcome. However, the original version (Caprini[orig:adm]) provided comparable predictability: the score of ≥6 predicted the unfavorable outcome with the sensitivity of 74% and specificity of 66%.

**Figure 5.**
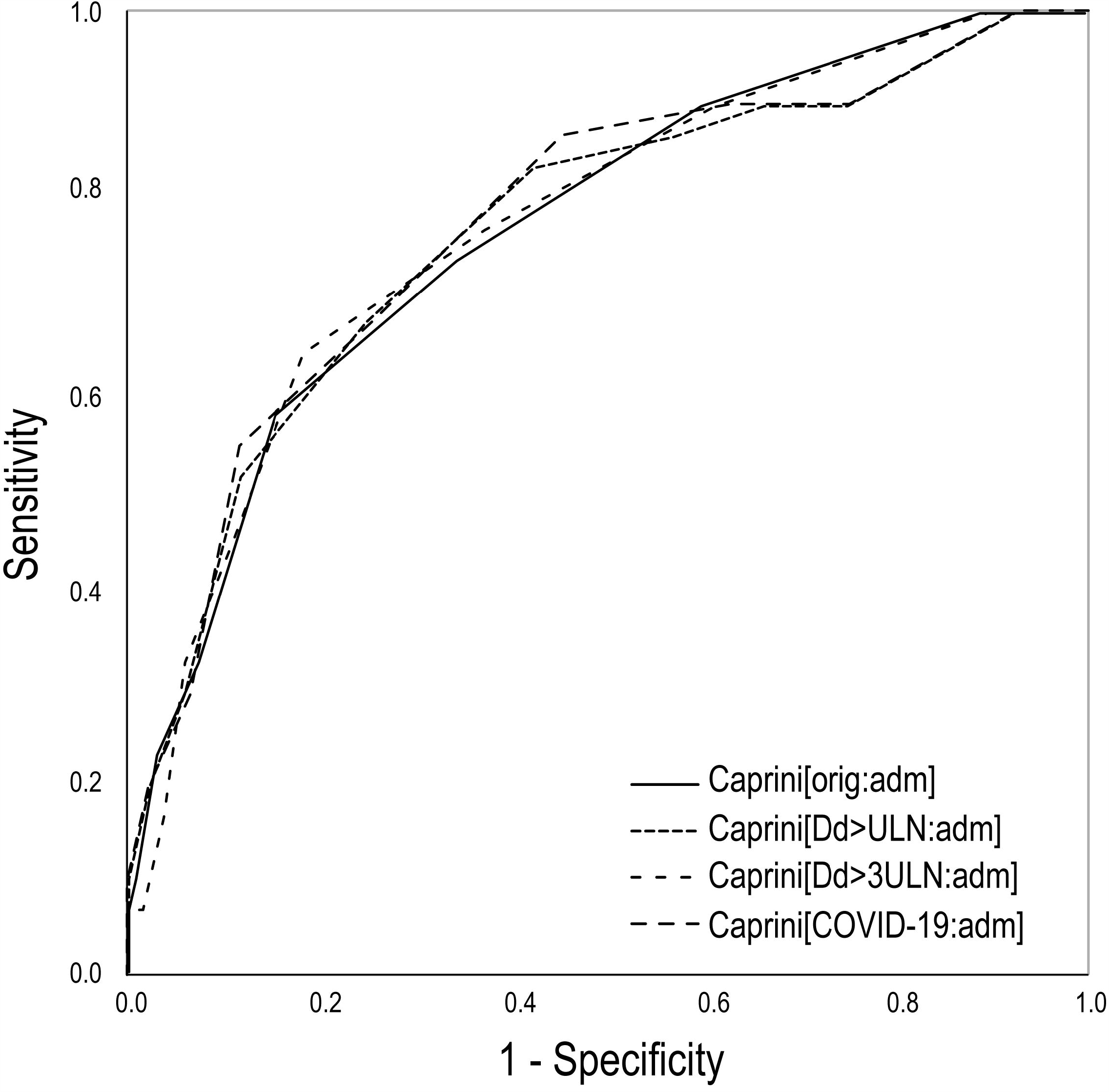
Predictability of different versions of Caprini score for unfavorable outcome in COVID-19 by the ROC curves. Unfavorable outcome combined symptomatic VTE, admission to the intensive care unit, the requirement for mechanical ventilation and death Caprini[orig:adm] – the original score assessed at admission; Caprini[Dd>ULN:adm] – considering 3 points if D-dimer increased over ULN, score assessed at admission; Caprini[Dd>3ULN:adm] - considering 3 points if D-dimer increased >3 times over ULN, score assessed at admission; Caprini[COVID-19:adm] – considering 2 points for asymptomatic infection, 3 points for symptomatic infection, 5 points for symptomatic infection with positive D-dimer, score assessed at admission; ROC – receiver operating characteristic

## Discussion

The Caprini score is the most validated risk assessment tool for VTE that was evaluated in about 5 million medical and surgical patients worldwide.^35-37^ The previous study showed improved predictability when the results of viscoelastic measurements supplemented the original score.^38^ However, it has not been validated in patients infected by SARS-CoV-2 to date. Despite this, the American Venous Forum has already recommended using the Caprini score for VTE risk assessment at admission and discharge.^19^ They referenced the original Caprini score in their recommendations. The score of ≥8 was suggested as an indication for the intensification of pharmacological prophylaxis in the hospital and extended prophylaxis after discharge. Our study is the first to use a COVID-19 modified Caprini score, and compare it to the original score. We also tested some theoretical score modifications based on the D-dimer level. At the same time, our study shows that an original score of ≥7, as calculated at admission, and a score of ≥11, as calculated at discharge, are associated with increased risk of VTE despite the administration of LMWH (predominantly intermediate doses). Such patients may be considered as an extremely high-risk group that requires special attention. The threshold of 11 points has been identified previously as a criterion for extremely high-risk of VTE in surgical patients.^24, 38^ The importance of the D-dimer level in COVID-19 was introduced recently. It appears to be associated with unfavorable outcomes, increased mortality, and VTE.^3, 8, 10, 27-34^ The thresholds for VTE verification were found at the levels of 1.0-2.3 mg/L^3, 8, 10^. The prophylactic doses of LMWH increased survival in patients with D-dimer level >6 times the ULN.^39^ The American Venous Forum proposed therapeutic anticoagulation for subjects with D-dimer >3 times ULN.^19^ However, in our study, the suggested level of D-dimer supplemented the scale did not improve the predictability of the original Caprini score for VTE. At the same time, considering any elevation of D-dimer improved the predictability and specificity of the original score when calculated at admission, but not at the discharge. In general, all evaluated models demonstrated good predictability with minor differences.

The predictive value for the unfavorable outcome was similar for all models with limited advantage of the one considering the new COVID-19 points. It was the only issue where the suggested modification appeared useful. However, the original version demonstrated sufficient predictability. Of note, any change of the original Caprini score affects the predictive threshold for extremely high-risk group, which could vary by 4-5 points. So the prognosis for VTE and detection of patients requiring intensified prophylaxis and/or additional imagining tests for VTE should be based on a specific threshold of every version of the scale (Table IV).

Interestingly, the incidence of VTE and mortality rate in our study appeared to be lower than previously reported for the general ward.^4, 7, 10^ This fact may be related to the use of intermediate to therapeutic doses of LMWH in 97,6% of all patients. There is a limited evidence that therapeutic anticoagulation can improve survival in COVID-19 patients requiring mechanical ventilation.^40^ However, the risk-benefit ratio of intensified anticoagulation should be confirmed in robust randomized clinical trials.

The study was limited by a retrospective character, small sample size, absence of the total instrumental screening for VTE, and lack of follow-up after discharge. Despite these disadvantages, it may be essential to improve VTE risk stratification in COVID-19 patients.

## Conclusion

The study identified a direct correlation between the Caprini score and the risk of VTE or unfavorable outcomes in COVID-19 patients. The score of ≥7, as calculated at the admission, or ≥11, as evaluated at the discharge, predicts symptomatic venous thromboembolism despite pharmacological prophylaxis. The predictability of the original score may be improved by the implementation of a D-dimer or specific COVID-19 points. However, the original Caprini score is essential. Further studies are needed to determine if there are any clinical benefits using the modified COVID-19 Caprini score. It will be especially important to track 90-day clinical events based both on the discharge modified Caprini score and the use of post-discharge prophylaxis.

## Data Availability

The data set is available by the request to the corresponding author.

